# Retinal aging transcriptome and cellular landscape in association with the progression of age-related macular degeneration

**DOI:** 10.1101/2022.04.03.22273375

**Authors:** Jiang-Hui Wang, Raymond C.B. Wong, Guei-Sheung Liu

## Abstract

Age is the main risk factor for age-related macular degeneration (AMD), a leading cause of blindness in the elderly, with limited therapeutic options. Here we systematically analyzed the transcriptomic characteristics and cellular landscape of the aging retina from controls and patients with AMD. We identify the aging genes in the retina that are associated with innate immune response and inflammation. Deconvolution analysis reveals that the estimated proportion of M2 and M0 macrophages is increased and decreased, respectively with both age and AMD severity. Moreover, we find that Müller glia are increased with age but not with disease severity. Several genes associated with both age and disease severity in AMD, particularly *C1s* and *MR1*, are strong positively correlated with the proportions of Müller glia. Our studies expand the genetic and cellular landscape of AMD and provide avenues for further studies on the relationship between age and AMD.

## Main

Aging is one of the most important demographic risk factors for age-related macular degeneration (AMD)^1, 2^, the leading causes of blindness among people aged over 55-year-old in the developed world^3^. A review summarizing the prevalence of AMD over age from various studies suggested a clear trend that the prevalence of AMD regardless mild or severe clinical features is gradually increased with age^2^. Advances in sequencing technologies, genetic analyses, such as genome-wide association studies (GWAS), using large numbers of samples from donors with AMD, enables identification of common and rare variation that is associated with AMD^4^. Moreover, expression quantitative trait loci (eQTL) analysis using transcriptional profiles of postmortem retinas from 453 controls and AMD patients expanded the genetic landscape of AMD^5^. Such studies also have implicated roles for the immune system, in particular the dysregulation of the complement system, in the pathogenesis and severity of AMD^6^. While these studies provided useful information on possible causal genes or immune cell types associated with AMD, these is little information on how age affects the progression of AMD.

In this study, we systematically assess the transcriptomic profiles, immune and retinal cellular landscapes of the aging retinas in association with the progression of AMD. We define the aging genes in the retina and show that the aging retina is transcriptionally featured by innate immune response and inflammation. Deconvolution analysis reveals increased and decreased proportion of M2 and M0 macrophages, respectively with both age and disease severity during the progression of AMD. We further identify specific genes that altered with age and disease severity, which are relevant to immune response, leukocyte activation and multicellular organismal homeostasis. Our results suggest that Müller glia are increased with age but not with disease severity, further indicating that age is the principal risk factor that uniquely contributes to the severity of AMD. Our study provides pinpointed detailed genetic and cellular characteristics of the aging retina that contribute to the severity of AMD.

## Results

We analysed the Eye Genotype Expression (EyeGEx) database, a public resource of retinal transcriptional profiles from patients with AMD^7^. A total of 453 previously quality-controlled^7^ retinal RNA-seq profiles from controls and cases of different ages (50-109 years old) and at distinct stages of AMD were analyzed (Fig.1a). The Minnesota Grading System (MGS) was used in this database to classify the disease severity. MGS1 categorizes donor retinas without AMD symptoms and serves as controls, whereas MGS2 to MGS4 categorize donor retinas at the progressively advanced AMD stages.

### Identification of age-associated genes in the human retina

Gene expression levels in the EyeGEx database are previously defined by a number of clinical variables, including MGS level, sex and age^7^. By controlling for MGS level and sex, therefore, we identified two clusters of genes associated with progressive aging (adjusted *p* <0.05, Supplementary Data 1) using a likelihood-ratio test^8^. Specifically, we found that expression levels of 445 and 57 genes were increased and decreased over age, respectively (Fig. 1b, hereafter referred to Age-up and Age-down genes, respectively). A heatmap of expression profile of all Age-up and Age-down genes also shows the progressive changes of these genes over age (Fig. 1c). Gene ontology (GO biological process) analysis of Age-up genes by Metascape, a comprehensive and combinational knowledgebase^9^, indicates that innate immune responses, leukocyte activation and inflammatory responses are among the top GO terms (Fig. 1d, Extended Data Fig. 1, Supplementary Data 2). Given the limited number of Age-down genes, there were no significant GO terms enriched. Our results were consistent with previous reports that retinal aging is a major risk factor of developing various retinal degenerative diseases mainly via dysregulation of innate immune systems, including AMD, diabetic retinopathy and glaucomatous retinopathy^10, 11^. Studies on the retinal immune cells in particular macrophages suggested its activation in the retina is reported in both AMD patients^12^ and old mice^13^.

**Fig. 1.**
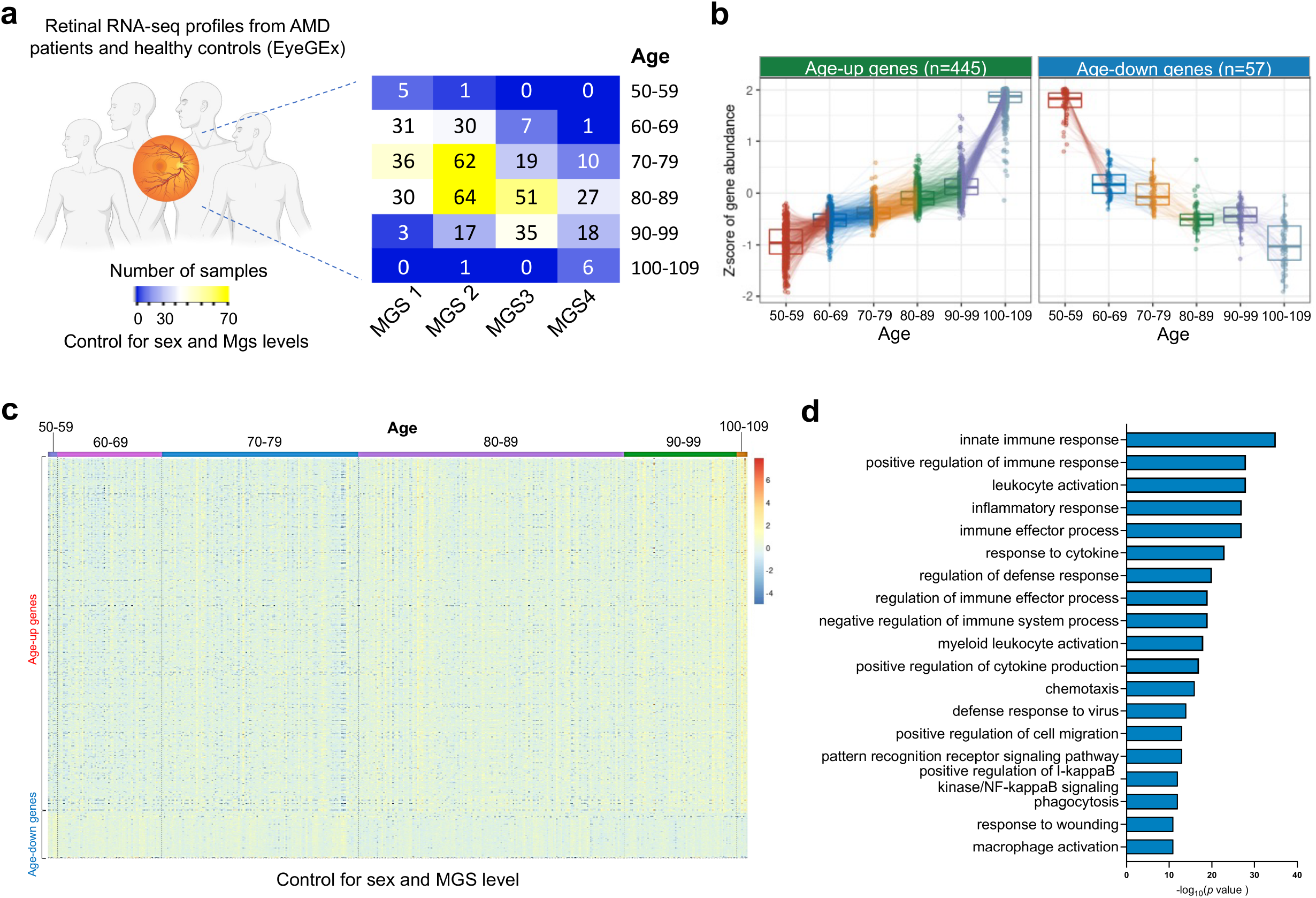
Identification of age-associated genes in the human retina from donors with AMD. **a**, Demographics of the human retina RNA-Seq profiles in the EyeGEx dataset (n=453, previously quality-controlled dataset), detailed by age group and AMD severity (MGS level). **b**, Tukey boxplots (interquartile range (IQR) boxes with 1.5× IQR whiskers) of age-associated genes in the human retina. Expression of Age-up genes increases with age (left, n=445), while expression of Age-down genes decreases with age (right, n=57). Age-associated genes were defined with DESeq2 two -sided likelihood-ratio test (adj. *p* < 0.05) by controlling for sex and. MGS level. Adjusted gene expressions were shown as z-score. **c**, A heatmap of expression level of all age-associated genes in all samples across different age group, adjusted for sex and MGS levels. **d** Functional enrichment analyses (biological processes) of age-up genes by Metascape.

### The immune cellular landscape in the retina of patients with AMD

In order to assess different immune cell types and to understand their relationship with age or disease severity (ranked by MGS level). in the retina of patients with AMD, we deconvoluted the EyeGEx database by CIBERSORTx, a machine learning method that enables inference of cell-type-specific gene expression profiles without physical cell isolation^14^. LM22, a signature matrix that enables distinction of 22 immune cell types, was used as a reference for deconvolution analysis^15^. We specifically looked at how age or MGS level as an individual factor impact the cellular proportions of immune cells using an ordinal logistic regression (OLR) model. By controlling for sex and MGS level, we found that proportions of M1 macrophages, M2 macrophages, monocytes were significantly increased with age, while M0 macrophages were decreased (Extended Data Figs. 2 and 3, Supplementary Data 3). By controlling for sex and age, interestingly, we observed that proportions of activated NK cells and M2 macrophages were significantly increased with MGS level, while M0 macrophages were also decreased (Extended Data Figs. 4 and 5, Supplementary Data 4). It is known that human peripheral monocytes were differentiated into inactivated M0 macrophages and then polarized to M1 and M2 phenotypes under various stress or stimuli^16^. Previous studies have shown that numbers of both M1 and M2 macrophages were increased in the retina^12^ or aqueous humors^17^ of a small number of neovascular AMD patients or laser-induced murine model with choroidal neovascularization^17, 18^. In line with these reports, our results further revealed that age alone as a predominant factor that triggers the polarization of M0 macrophages towards to pro-inflammatory M1 and pro-angiogenic M2 phenotypes^12^, while disease severity alone seems majorly affecting the numbers of M2 macrophages that feature in the pro-angiogenic process. Collectively, these results together showed that activated M2 macrophages caused by age and disease severity is the major immune cell type involved in the immune response in the pathogenesis of AMD.

### Identification of age- and disease-severity-associated genes in relation to progression of AMD

We next investigated whether retinal aging is related to MGS level in AMD patients. Using the OLR model that controls sex and age, we correlated age-associated genes to MGS level, resulting in 50 genes (hereafter referred to Age-MGS genes, *p* <0.05) that their expression levels were significantly altered with the change of MGS level (Fig. 2, Extended Data Fig. 6, Supplementary Data 5). Among these Age-MGS genes, expression levels of 7 genes (including *CYP26B1, DHRS3, MATN1, RPL15, AL136454*.*1, CXCL11, and CYP26A1*) were reduced with disease severity, while expression levels of the rest of the genes were escalated with disease severity. Interestingly, none of the identified Age-MGS genes were previously reported as AMD-associated genes in genome association studies. We then explored the biological processes of each Age-MGS gene involved by analyzing their corresponding Gene Ontology (GO) terms (Supplementary Data 6). We found that nearly half of Age-MGS genes are involved in the activation of immune cells or inflammatory pathways (Supplementary Data 6), indicating a strong immune response and inflammation occurred during the development of AMD over increased age. Other Age-MGS were clustered to groups with other biological processes, such as retinal endothelium-mediated angiogenesis and retinoic acid metabolic process.

**Fig. 2.**
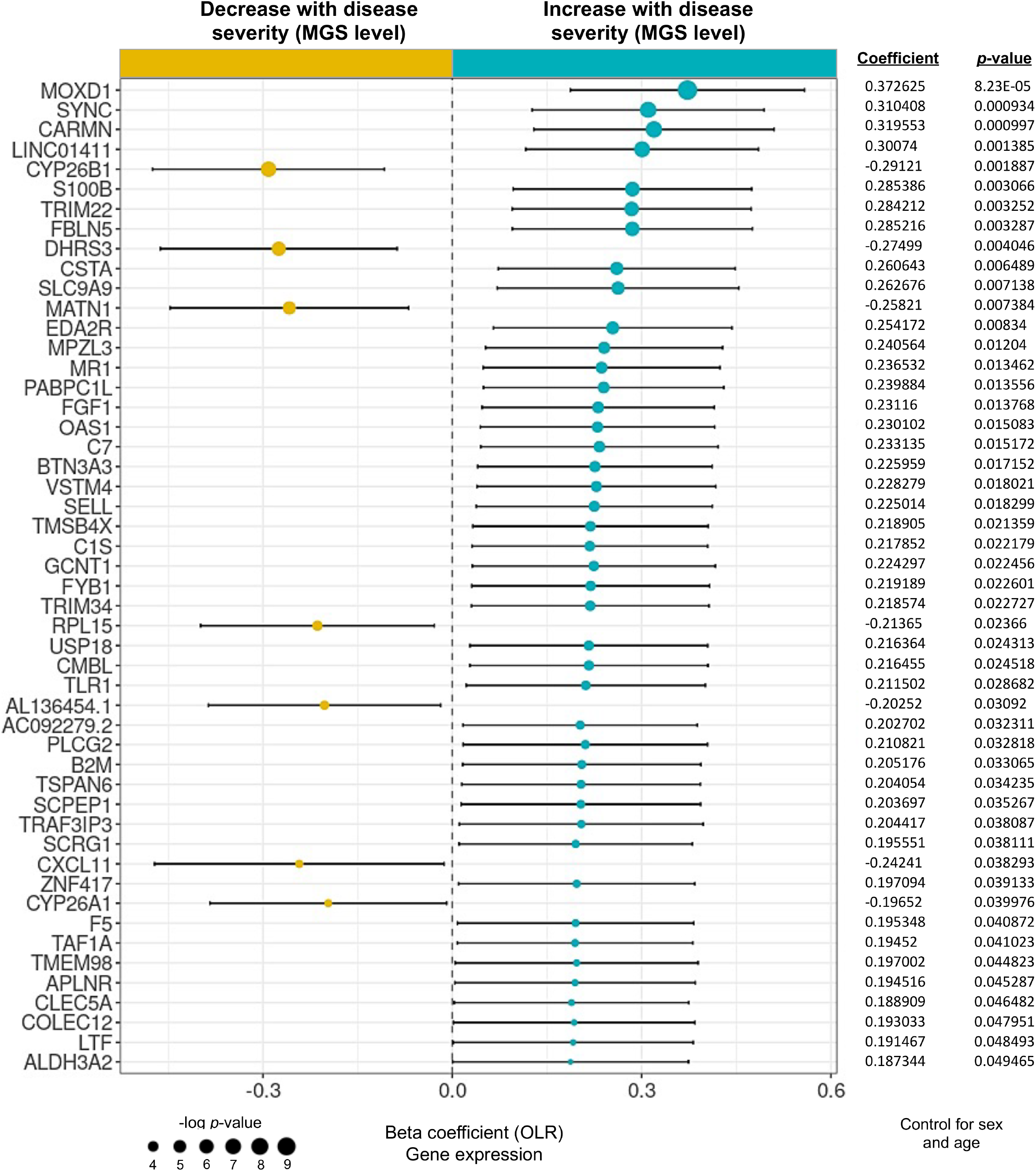
Identification of age-associated genes progressively altered with AMD severity (MGS level). Forest plot of age-associated gene having significant correlation with MGS level (*p* < 0.05, n=50). Positive coefficients indicate age-associated genes increase in expression with MGS level, while negative coefficients indicate age-associated genes decrease in expression with MGS level (i.e. Age-MGS genes). Statistical analyses of MGS association for age-associated genes were assessed by a nonparametric ordinal logistic regression model that controls for sex and age. Point sizes are scaled by statistical significance. Error bars represent 95% confidence intervals.

Although none of the Age-MGS genes we identified has been previously reported in genome association studies in AMD, many plays critical roles in immune responses and inflammation in the retina. For example, S100 calcium binding protein B (*S100B*), a well-known biomarker of active neural distress that is involved in many neurodegenerative diseases^19^, are pro-inflammation and can induce retinal degeneration in animal models^20, 21, 22^. L-selectin (*SELL*), a type-I transmembrane glycoprotein and leukocyte-expressed cell adhesion molecule^23^, mediates retinal inflammation by selective recruitment of leukocytes subsets, triggering adhesion, diapedesis and migration of inflammatory cells^24^.

Retinal endothelium plays important roles in maintenance of retinal statics particularly in the development of retinal vasculature. Among 50 Age-MGS genes, a cluster of genes are related to endothelium or angiogenesis (Supplementary Data 6), including apelin receptor (*APLNR*), fibroblast growth factor 1(*FGF1*), matrilin 1 (*MATN1*), V-set and transmembrane domain containing 4 (*VSTM4*), fibulin 5 (*FBLN5*), myelin protein zero like 3 (*MPZL3*), and serine carboxypeptidase 1 (*SCPEP1*), and their expression (except for *MATN1*) were increased with the disease severity, indicating the involvement of endothelial cells throughout the different stage of AMD. *APLNR* expression is limited in the veins and proliferative endothelial cells during physiological retinal angiogenesis^25^, suggesting endothelial cells might be active at the early stage of AMD. Indeed, using APLNR inhibitors can selectively prevent pathological retinal angiogenesis in an oxygen-induced retinopathy mouse model^25^. Studies have shown FGF1 plays a role in retinal neovascularization through organization of various angiogenic pathways and coordination of cell-cell interactions^26, 27^. We found that *MATN1* expression was decreased with the disease severity. This result is consistent with a study that showed increased angiogenesis during fracture healing in a *MATN1* deficient mouse, and purified MATN1 proteins enabled inhibition of angiogenesis both *in vitro* and *in vivo*^*28*^. Of note, *VSTM4, FBLN5, MPZL3* and *SCPEP1*, have been reported to promote sprouting angiogenesis by endothelial cells^29, 30, 31, 32^, while their role in retinal angiogenesis remains unstudied.

We also found that a group of Age-MGS genes are related to the regulation of retinoic acid metabolic process (Supplementary Data 6), including cytochrome P450 family 26 subfamily A member 1 (*CYP26A1*) or B member 1 (*CYP26B1*), dehydrogenase/reductase 3 (*DHRS3*) and serine carboxypeptidase 1 (*SCPEP1*), and their expression (except for *SCPEP1*) were reduced with disease severity. Retinoic acid is an active metabolite of vitamin A (retinol), an essential signaling molecule involved in the normal development of the ventral retina and optic nerve^33^ as well as maintenance of the blood-retinal barrier^34^. Dysregulation of retinoic acid can cause degenerative retinal diseases. *CYP26A1*, a retinoic acid catabolizing enzyme, was reported to be enriched in the fovea in the human retinas and functions for photoreception in rod-free zone generation^35^. Reduction of *CYP26A1* may affect degradation of retinoic acid, ultimately causing dysfunctionality of photoreceptors. Similarly, *CYP26B1* is also responsible for retinoic acid metabolism^36^, however, its role in the retina has not been investigated yet. *DHRS3*, a retinal reductase and a possible gene target of all-trans-retinoic acid, has been reported to be reduced in the liver during inflammation likely resulting in the perturbation of the whole-body vitamin A metabolism that occurred in conditions with inflammatory stress^37^. Together, these results imply that reduction of genes associated with retinoic acid-metabolism may disrupt functionality in photoreceptors, a typical feature seen in AMD patients^38^.

Some Age-MGS genes were not clustered into the three groups above mentioned (Supplementary Data 6) but their significance in relation to disease severity remained high (Fig. 2). Few studies investigated function of Monooxygenase DBH like 1 (*MOXD1*), however, GO annotation of *MOXD1* revealed its involvement in octopamine biosynthetic process, dopamine catabolic process, and norepinephrine biosynthetic process (Supplementary Data 6). Our results showed an increased expression level of *MOXD1* with disease severity, implying a progressive elevation of those neurotransmitters in the retina of AMD patients with age and disease severity. Increasing evidence showed that neural inputs, such as chronic stress or inflammation, resulted in the release of neurotransmitters (e.g. norepinephrine) at specific vessels in the blood-brain barrier, that in turn augment the expression of chemokines in the endothelium to establish gateways through which immune cells can reach the central nervous system^39, 40, 41, 42^. Given that the retina displays similarities to the brain and spinal cord in terms of functional, anatomic and immunological properties^43^, it is possible that neurotransmitters in the retina might be also increased to stimulate immune responses under chronic stress resulting from AMD. Type III intermediate filament proteins including glial fibrillary acidic protein (*GFAP*), vimentin, desmin, peripherin and syncoilin (*SYNC*, ranked No. 2 in the Age-MGS gene list according to *p* value, Fig. 2), have been reported in association with retinal gliosis^44^, a feature of AMD. A quantitative study in particular investigated the roe of *GFAP* expression at early and late stage of AMD and showed that upregulation of *GFAP* in the inner retina was predominantly related to drusen, while increased expression of *GFAP* in outer retina was associated with disruption of the RPE and blood-retinal barrier^45^. This result suggested that other type III intermediate filament proteins, such as SYNC, may be involved in the gliotic response in the retina subjected to chronic stress.

### Correlation of Age-MGS genes and their enrichment in retinal cell types

We next sought to understand the correlation between the expression of Age-MGS genes. Correlation analysis revealed 3 clear clusters of Age-MGS genes with correlated expression using the EyeGEx database with sex and age controlled. (Fig. 3a). For ease of description, these clusters were numbered as cluster A, B and C. Cluster A consists of zinc finger protein 417 (*ZNF417), AC092279*.*2, MPZL3* and TATA-box binding protein associated factor, RNA polymerase I subunit A (*TAF1A*), most of which are relevant to regulation of transcription or gene expression (Supplementary Data 6). Cluster B contains 38 Age-MGS genes that are involved in various biological pathways. Of which, complement C1s (*C1s*), major histocompatibility complex (MHC) class I-related (*MR1*) and solute carrier family 9 member A9 (*SLC9A9*) have stronger correlation (|R| ≥0.4) with other genes in this cluster, suggesting their potential role in coordinating other genes for multiple biological processes in AMD. *C1s* is a single chain glycoprotein and cleaves the components *C4* and *C2* to initiate the complement cascade when it becomes activated^46^. Transcriptome profiling of retina tissue from AMD donor eyes (31 normal and 26 AMD) suggested a direct role of dysregulation of *C1s* among other complement pathways in retinal degeneration^47^. Moreover, upregulation of *C1s* along with *C4a* and *C2* were found in retinas of rats subjected to photo-oxidative damage^48^. However, the functional role of *C1s* in retinal degeneration remains unclear. *MR1* molecule is distinct from MHC class I molecules that can present microbial riboflavin metabolites to a specialized subset of T cells known as mucosal-associated invariant T (MAIT) cells^49^. Interestingly, a study showed that *MR1* was up-regulated at early stages of retinal degeneration in DBA/2J mice featured an immune response in the iris and death of retinal ganglion cells^50^, suggesting its potential role in the progression of immune-relevant retinal diseases. *SLC9A9* is highly expressed in the brain and is implicated with immune regulation and endocytosis via mTOR signaling and cell survival^51^. However, little is known about its role in the retina. Cluster C consists of all down-regulated genes with disease severity, including *MATN1, RPL15, CYP26A1, CYP26B1 and DHRS3*, which are associated to regulation of retinoid metabolic process, extracellular matrix organization, and cytoplasmic translation (Supplementary Data 6). It is unclear how these molecules interact in the progression of AMD.

**Fig. 3.**
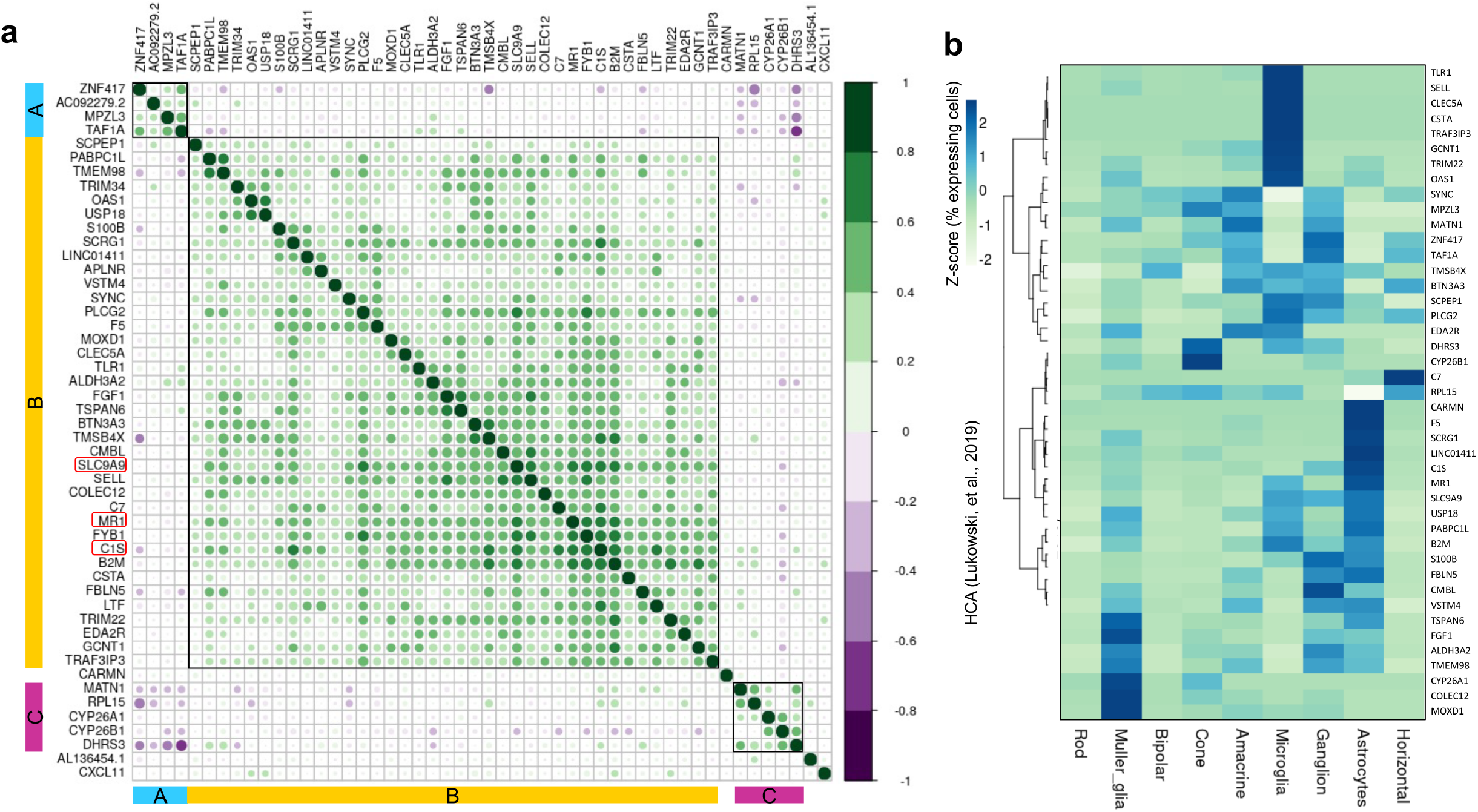
Correlation of Age-MGS genes and their enrichment in retinal cell types. **a**, Heatmap of the correlation matrix across 50 Age-MGS genes identified in Fig.2. Pearson’s correlation was calculated among 50 Age-MGS genes to show the co-expression patterns of genes in the heatmap. Colour key denotes the Pearson’s correlation higher than absolute value 0.3 between genes. There are 3 clear clusters (labelled as A, B and C) of Age-MGS genes with correlated expression adjusted by sex and age. Genes highlighted in red indicates their stronger correlation with many other genes. **b**, Heatmap showing the percentage of retina cells expressing each of Age-MGS genes, scaled by gene across the different cell types. Data are from the Human Cell Atlas (HCA).

To assess the cell type-specific expression pattern of the age-associated genes and Age-MGS genes, we further analyzed the neural retina single-cell RNA-seq (scRNA-seq) data from Human Cell Atlas (HCA, Extended Data Fig. 7)^52^. We found that the expression pattern of the age-associated genes is distinctive among 9 retinal cell types, and a higher percentage of cells expressing these genes are Müller glia, microglia, ganglion and astrocytes (Extended Data Fig. 8). Age-MGS genes are more enriched in Müller glia, microglia and astrocytes (Fig. 3b). Müller glia and astrocytes express a few similar Age-MGS genes but their percentage of expressing cells are clearly different (Fig. 3b). Consistent with our finding, a previous study reported that Müller glia and astrocytes among other major retinal cell types express the highest number of AMD-associated genes identified by a GWAS study^53^. Müller glia and astrocytes assist metabolism of retinal neurons through releasing trophic factors, recycling neurotransmitter glutamate, and controlling extracellular ion homeostasis^54, 55^, while astrocytes are resident macrophages and are implicated in neuroinflammatory changes during age-dependent retinal diseases, including AMD^56^. Together, these analyses highlight the cellular landscape of Age-MGS genes in the context of retinal cell types in which these genes are normally expressed. Of note, many Age-MGS genes were not expressed in any of the neural retina cell types, and their expression in retinal cell types in the condition of AMD is not clear. We also found that correlation between AMD-associated genes identified through different methodologies, such as LRT combined with OLR (present study), transcriptome-wide association study (TWAS using EyeGEx)^7^ and GWAS^4^, is clearly distinct (Extended Data Fig. 9a), indicating that these is limited interaction between these genes. However, genes identified by TWAS (9 genes) and GWAS (25 leading genes from 34 risk loci associated with AMD, sample size >15,000) are closer to each other as analyses of TWAS are based on GWAS result. Using HCA data, we validated leading AMD-associated genes previously identified GWAS^4^ and showed similar results that higher percentage of associated cells expressing AMD-associated genes are Müller glia and astrocytes (Extended Data Fig. 9b).

### Functional roles of Age-MGS-associated genes in AMD

To determine the functional roles of these Age-MGS genes involved in the pathogenesis of AMD, we applied functional enrichment analysis (Biological Process) by Metascape. The results revealed significant enrichment most related to regulation of innate immune response (Fig. 4a, Supplementary Data 7). Among which, innate immune response, leukocyte activation, and multicellular organismal homeostasis are centered in the cluster of GO terms and have most interaction (n ≥5) with other GO terms (Fig. 4b), suggesting their important roles in regulation of Age-MGS genes. Therefore, we combined genes involved in these biological processes (Supplementary Data 7) and focused on the 8 Age-MGS genes with significant changes with both MGS levels (Fig 4c) and age (Fig 4d)), including *C1s, C7, MR1*, phospholipase C gamma 2 (*PLCG2*), tripartite motif containing 22 (*TRIM22*), transmembrane protein 98 (*TMEM98*), *CYP26B1*, and *VSTM4*.

**Fig. 4.**
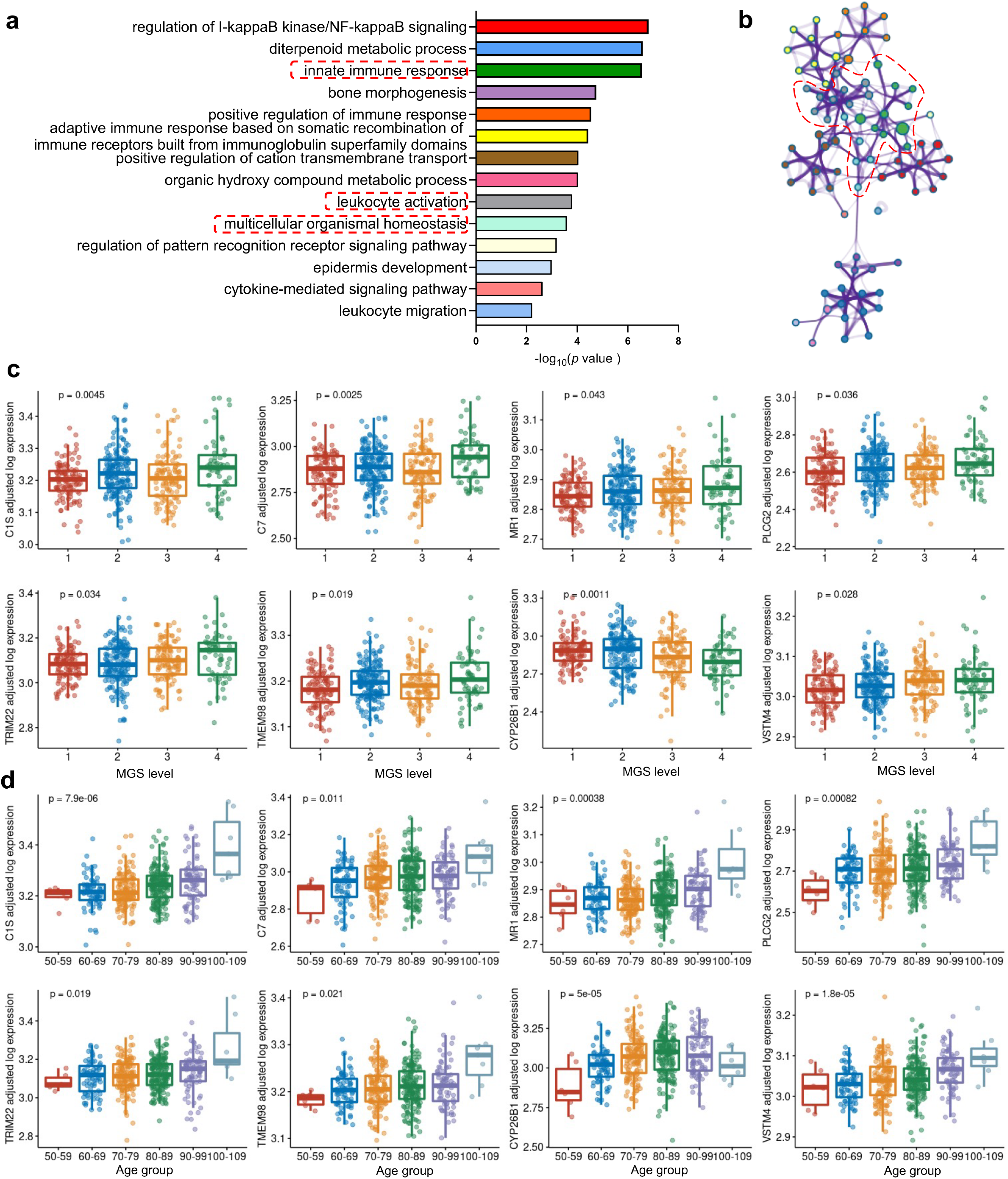
Functional roles of Age-MGS genes in AMD. **a**, Functional enrichment analyses (gene ontology annotations of biological processes) of 50 Age-MGS genes by Metascape. **b**, Interaction network of gene ontology annotations described in Fig. 4a. Annotations highlighted in red indicates that the specific biological process has the most interactions with other biological processes. **c**, Tukey boxplots (interquartile range (IQR) boxes with 1.5× IQR whiskers) showing the expression of *C1s, C7, MR1, PLCG2, TRIM22, TMEM98, CYP26B1*, and *VSTM4*. Gene expression value are shown as log-transformed, controlled for sex and age (Fig. 4c) or sex and MGS level (Fig. 4d). Statistical significance of age-associated or MGS-associated difference was assessed by two-sided Kruskal-Wallis test on the adjusted expression values.

*C1s* and *MR1* may play a central role in the regulation of innate immune responses because these genes have higher correlation scores with more Age-MGS genes that we aforementioned (Fig. 3a) and are reported to be related to retinal degeneration^48, 50^. However, their individual role in regulating immune response in the retina remains unclear. Complement *C7* as one of terminal proteins of complement pathway including *C5, C6, C8* and *C9* involves complement-mediated inflammation in the retina in AMD^57^ and is protective for AMD development in Caucasians^58^. These findings provided further evidence for complement overactivation in AMD pathogenesis^59^. An integrating analysis using *in silico* genetic and biological pathway data showed that *PLCG2* is associated with AMD based solely on genetic burden^60^. Our study in line with this finding showed a progressively increased expression level of *PLCG2* in the retina of AMD patients with age and MGS level. Both *TRIM22* and *TMEM98* are involved in biological processes such as immune response or inflammation (Supplementary Data 6), while their function in the stressed retina is not clear. *CYP26B1* as one of few down-regulated aging genes with disease severity play a role in regulation of retinoid metabolic process, while *VSTM4* contributes to the stability of retinal vasculature (Supplementary Data 6). Together, our results suggested that these identified 8 Age-MGS genes collectively regulate innate immune response, promote leukocyte activation and endothelial cell proliferation and ultimately retinal angiogenesis.

### The cellular landscape of the retina of patients with AMD and its relation to Age-MGS-associated genes

Previous studies have shown cellular changes occurred in the retina of patients with AMD, including Müller glia, astrocytes, microglia and retinal pigment epithelium^45, 61^. However, detailed quantification of cellular changes remains a challenge. Here we deconvoluted the bulk retina EyeGEx transcriptomes with CIBERSORTx to quantify cellular changes, using the scRNA-seq data from HCA as a reference (Extended Data Fig. 7, Supplementary Data 8). To increase the confidence of down-stream analyses, the cell types were retained if their estimated proportions were >0 in all samples (Extended Data Fig. 10).

To assess whether age as a risk factor affects cell type proportions, we used an OLR model to control for sex and MGS level and identified age-associated alteration in the proportions of retinal cell types. We observed that the proportions of Müller glia were significantly increased with age, while the proportions of horizontal and rod cells were significantly decreased with age (Fig. 5a, Extended Data Fig. 11, Supplementary Data 9). Next, we focused on the risk factor of MGS level to assess its effect on cell proportions by controlling for sex and age using an OLR model. Although the proportions of Müller glia and rod were increased and decreased, respectively, with MGS levels, the changes were not significant (Fig. 5b, Extended Data Fig. 12, Supplementary Data 10), suggesting that disease severity has limited effects to alter cell proportions in the retina of patients with AMD. Of note, ganglion cells were removed in Fig. 5b due to its zero estimated expression profile across all samples after adjustment for sex and age. Previously study using immunolabelling of glial fibrillary acidic protein (GFAP), a marker of retinal glial cells, observed morphological changes and upmodulation of astrocytes and Müller glia in the inner and outer retina, respectively in a small number of human donor retinas (normal=6, AMD=11)^45^. Rods loss in ageing and age-related maculopathy has been widely reported^62, 63^. Moreover, Rods die in older eyes without evidence of overt retinal pigment epithelial degeneration^62^, indicating that age is a main factor to cause rods loss. Horizontal cells form triad synapses with photoreceptors and bipolar cells to module signal transmission in between these cells^64^. scRNA-Seq profiling of the aging human retina revealed a horizontal cell subtype (ISL1^+^ CALB1^+^) were greatly reduced particularly in fovea, suggesting that age may affect different regions in the retina^65^. Together, these results suggested that age is a central risk factor in AMD that enables alterations in the proportions of cell types in the retina, i.e. Müller glia, horizontal cells and rod photoreceptors.

**Fig. 5.**
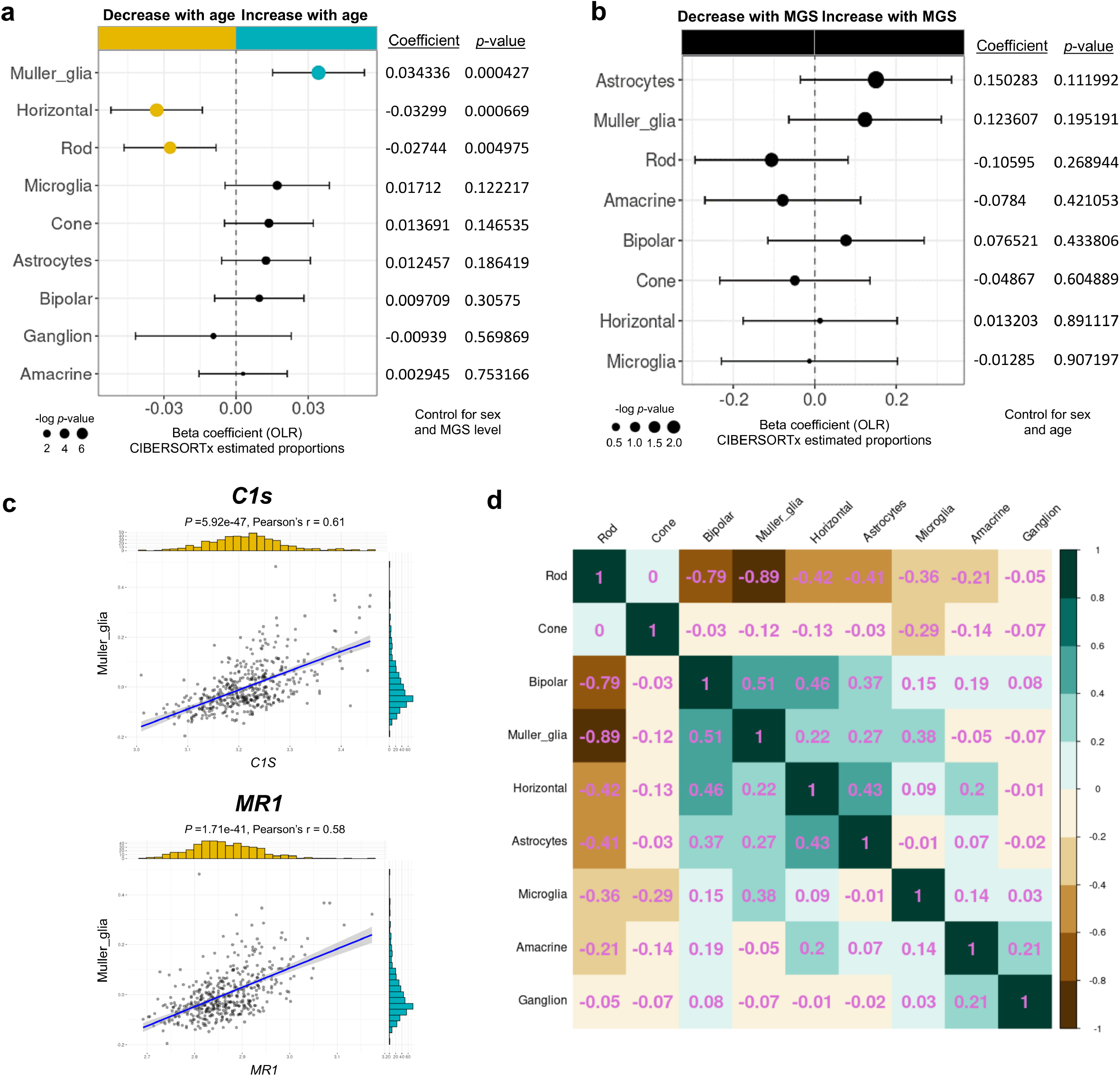
The cellular landscape of the retina of patients with AMD and its relation to Age-MGS genes. **a, b**, Forest plot of estimated proportions of retinal cells having significant correlation with age or MGS level, respectively (p < 0.05). Positive coefficients indicate the proportion of cells increases with age or MGS level, while negative coefficients indicate the proportion of cells decrease with age or MGS level. Statistical analyses of age association or MGS association for the proportions of cells were assessed by a nonparametric ordinal logistic regression model, adjusted for sex and MGS level (Fig. 5a) or sex and age (Fig. 5b), respectively. Point sizes are scaled by statistical significance. Error bars represent 95% confidence intervals. **c**, Analyses of the correlation between expression level of *C1s* or *MR1* and the estimated proportion of Müller glia, adjusted for sex and age. Pearson’s correlation coefficient was used to test the strength of linear relationships between gene expression and the estimated proportion of Müller glia. Black dots denote individual samples (n=453). Blue lines denote regression lines. **d**, Heatmap of the correlation matrix among retinal cell types, based on the proportions of retina cells estimated by CIBERSORTx and adjusted for sex and age. Pearson’s correlation was calculated among retinal cells to show the co-expression patterns of cells in the heatmap. Values in the heatmap indicates the Pearson correlation coefficient between two cell types.

As most of Age-MGS genes are enriched in retinal glial cells including Müller glia and the increased proportions of Müller glia with age, we specifically investigated the relationship between Müller glia and 2 core Age-MGS genes we identified (Fig. 5c). We showed that expression of *C1s* and *MR1* have a strong positive (Pearson correlation coefficient, 0.5< r <1) relationship with the estimated proportions of Müller glia. We also observed strong negative (−0.5 < r <-1) correlation between expression of *C1s, MR1* and the estimated proportions of rod photoreceptors (Extended Data Fig. 13a), while no significant or weak positive correlation (0< r <0.3) between these two genes and the estimated proportions of horizontal cells (Extended Data Fig. 13b). These results suggested that *C1s* and *MR1* are potential core Age-MGS genes to regulate Müller glia and rod photoreceptors but not horizontal cells that are involved in retina aging. Notably, among the three cell types there is a relatively higher percentage of Müller glia expressing *C1s* and *MR1* (Fig. 3b). Photoceptor degeneration and Müller glia activation are indicative of structure and molecular abnormalities in the development of AMD^66^, suggesting their possible complementary relationship in this form of retinal degeneration. Indeed, our results revealed that there is a strong negative correlation (r = -0.89) between rod photoreceptors and Müller glia based on its estimated proportions after controlling for sex and age (Fig. 5d).

## Discussion

In the present study, we systematically analyzed the retinal aging transcriptome and cellular landscape in association with the progression of age-related macular degeneration. We revealed that the aging retina is characterized by a range of biological processes, in particular innate immune response and inflammation that could lead to the progressively severe AMD. We specifically studied the immune cell profiles of the EyeGEx database and identified the upregulation of M2 macrophages and downregulation of M0 cells with both age and disease severity in AMD patients. We subsequently pinpointed a group of retinal aging genes that change with disease severity (Age-MGS genes) by controlling for sex and age. Many of the Age-MGS genes have function mainly in relation to immune response, endothelium-mediated angiogenesis and retinoic acid metabolic process. Through gene correlation analysis, we demonstrated that *C1s, MR1* and *SLC9A9* have stronger correlation with other Age-MGS genes, and all are involved in regulation of immune response. By integrating these data with the single-cell transcriptome of the human neural retina, we further identified specific cell types that normally express these Age-MGS genes. GO enrichment analysis of the Age-MGS genes further suggested the aging and AMD retina is characterized by innate immune response, leukocyte activation and multicellular organismal homeostasis. We further identified 8 Age-MGS genes in these biological processes that potentially play critical roles in the progress of AMD development. Through bulk deconvolution analyses, we showed that the proportions of Müller glia are increased with age, while the proportions of rod photoreceptors and horizontal cells are decreased with age. Moreover, the proportions of all retinal cell types do not have significant changes with disease severity after factoring out sex and age, suggesting that age is potentially the central risk factor that uniquely contributes to the severity of AMD. Correlation analysis between 8 Age-MGS genes and the proportions of cells with significant alteration revealed that *C1s, MR1* and *TMEM98* have a strong relationship with Müller glia and rod photoreceptors, both of which are negatively correlated. Future research to investigate the function of these Age-MGS genes in the retina would be important to understand the mechanism underly AMD pathology.

Since the transcriptome data from human donors are complexed with a variety of latent factors, such as sex, age and clinical diagnosis, it is important to use batch correction to adjust those factors for the downstream analysis. Throughout our analysis, all the raw data were subjected to the batch correction before relevant analysis. We noted a study using various deconvolution algorithms to analyze involvement of retinal cell types in AMD patients with different databases, including EyeGEx^67^. However, all the raw data from this study was not subjected to batch correction, thus the conclusion resulted from was less stringent compared to the present study. A clear example of difference between adjusted and unadjusted data analysis can be referred to the cell type proportion estimated by CIBERSORTx (Extended Data Fig. 3, 5, 11 and 12). There were several limitations of this study. For examples, the sample size in each of age group is relatively small particularly the group of 100-109, which likely affects the number of aging genes defined by the LRT model. Moreover, the HCA dataset does not include RPE cells for which we might miss information on how the Age-MGS genes involving in the degeneration of RPE cells in AMD^68^. In addition, we used the human hematopoietic immune cell matrix, LM22, to deconvolute immune cell profiles in the retina, potentially missing information on resident immune cell types, such as microglia, in neural tissues including brain and retina. Future studies to determine the change of the expression of Age-MGS gene in RPE cells and to generate transcriptome dataset for resident immune cells in neural tissues would be helpful. Our study defined the aging genes in the retina using the LRT model by controlling for sex and disease severity of AMD and identified novel ageing genes associated with AMD progression. We also highlighted the potential for using deconvolution analysis to understand cellular and transcriptomic changes in AMD. The functions of most of the Age-MGS genes remain unclear in the retina and more research is needed to understand their role in progressive development of AMD. Our analyses provide potential directions for subsequent research on the aging retina for which AMD progressively gets worse over age.

## Methods

### Data accession

RNA-Seq raw counts of the Eye Genotype Expression (EyeGEx) database were obtained from Gene Expression Omnibus (GSE115828, accessed on 11^th^ January 2022)^7^. Raw counts were converted to transcripts per million (TPM) in R (v4.1.0) for downstream analyses. Quality-controlled samples (105 MGS1, 175 MGS2, 112 MGS3, and 61 MGS4 samples) were selected from the raw dataset using the subset function from the R package Seurat^69^ (v4.0.5). Detailed clinical information including patient sex, age and MGS level were accessed from GSE115828. Single-cell transcriptomes of human neural retina were directly downloaded from Human Cell Atlas (https://www.humancellatlas.org/, E-MTAB-7316, accessed on 15^th^ January 2022)^52^.

### Identification of age-associated genes in human neural retina from donors with AMD

To identify age-associated genes, the RNA-Seq raw count from quality-controlled samples was analyzed by DESeq2^8^(v1.32.0), using the likelihood-ratio test (LRT). According to the design of batch correction in EyeGEx database, gene expression was determined by three variables, including sex, age and MGS level^7^.Thus, sex, age and MGS level were included in the LRT model for controlling these factors. Age was categorized into decades, including 50-59, 60-69, 70-79, 80-89, 90-99, and 100-109, for downstream analyses. Significant age-associated genes were determined by adjusted *p* < 0.05. The resulting genes were then scaled to z-score and clustered using the degPatterns function from the R package DEGreport (v1.28.0). Gene clusters with consistent and progressive changes (increasing or decreasing with age) were selected for downstream analyses.

Enrichment analyses for selected genes were performed using Metascape (https://metascape.org/gp/index.html#/main/step1), a web-based tool that involves a variety of terms including GO Biological Processes^9^. Importantly, Metascape provides more frequently updated bioinformatics analyses than DAVID^70^.

### Identification of age-MGS-associated genes in human neural retina from donors with AMD

To identify age-MGS associated genes, the TPM values of age-associated genes from both clusters (increasing or decreasing with age) were analyzed by R package MASS (v7.3-54) and ordinal (v2019.12-10) using ordinal logistic regression (OLR) model^71^. Using Cumulative Link Model function in ordinal^71^, we controlled sex and age to screen input genes that are associated with MGS level. Age-MGS-associated genes were determined at a significance threshold of *p* < 0.05. The resulting genes were represented in a forest plot and ranked according to its beta coefficient.

For gene-gene correlation of Age-MGS genes, the raw counts were processed by Variance Stabilizing Transformation function in DESeq2^8^, followed by statistical adjustment for sex and age using the Remove Batch Effect function in limma^72^ (v3.48.3). The correlation heatmap was plotted using R package corrplot^73^ (v0.92).

### RNA-Seq gene expression visualization and statistical analyses

For visualization of adjusted gene expression levels, the raw counts were processed by Variance Stabilizing Transformation function in DESeq2^8^, followed by statistical adjustment for sex and MGS level or sex and age using the Remove Batch Effect function in limma^72^ (v3.48.3). Comparison of gene expression among different age groups or MGS groups are presented as Tukey boxplots, with interquartile range boxes and 1.5x interquartile range whiskers. Two-tailed Mann-Whitney test was used to assess pairwise comparisons in the plots, while Kruskal-Wallis test was used to perform statistical comparison across all the age groups or MGS groups. Such statistics were performed to confirm the results of age-associated genes generated by the two-sided DESeq2 LRT^8^.

### Normal human neural retina scRNA-Seq data analysis

scRNA-seq raw counts from HCA were analyzed using Seurat^69^ (v4.0.5). Cells with >200 detected genes and expressing < 10% mitochondrial genes were retained, resulting in 19,950 cells. Genes were retained if they were expressed in ≥ cells. The LogNormalize method was used to normalise the dataset, followed by removing inherent variation caused by mitochondrial gene expression and the number of unique molecular identifiers per cell^52^. Using the graph-based shared nearest neighbour method, clustering at a resolution of 0.9 was performed on PCA-reduced expression data because it yields high maximum cluster assignment probability for most of the cells. Clusters were visualized using uniform manifold approximation and projection (UMAP). Retinal cell markers were referred to studies analyzing retina scRNA-Seq data^52, 53^.

Of the 50 Age-MGS-associated genes identified from the EyeGEx transcriptome, 43 genes were matched in the HCA dataset. To calculate the proportions of cells expressing a given gene, the expression matrices were converted to binary matrices using the threshold of expression > 0^74^. Expression frequencies of a given gene in different cell types were determined as per the cell type annotation resulting from above analyses. We further scaled the expression frequencies in cell types in z-score using the NMF package (v0.23.0).

### Deconvolution analyses of the immune cells and retinal cells from donors with AMD

To infer the estimated proportions of infiltrating immune cells or retinal cells of each bulk retinal transcriptome, we used the CIBERSORTx algorithm, a deconvolution algorithm that uses a set reference gene expression value^14^. For the immune cellular profile, we used the signature matrix, LM22, defining 22 infiltrating cells^75^. Normalized data (TPM value) of the EyeGEx transcriptome were uploaded to the CIBERSORTx web portal (https://cibersortx.stanford.edu/), with the algorithm run using LM22 signature matrix at 100 permutations with B-mode batch correction.

For retinal cellular profile, we derived a signature matrix using HCA scRNA-Seq dataset. 10,000 retinal cells including rod, Müller glia, bipolar, cone, amacrine, microglia, ganglion, astrocytes, and horizontal cells, were acquired using the Seurat subset function and labelled with corresponding cluster identities. The labelled cells were used to establish a single-cell signature matrix with the following settings: Min.Expression (0.05), replicates (10), and sampling (0.5), kappa (999), q-value (0.01) and No. Barcode Genes (100-500). The resulting signature matrix was used to impute retinal cellular fraction from the EyeGEx transcriptome with S-mode batch correction. The number of permutations to test for significance were set at 100. Statistical significance of age- or MGS-level-association was assessed by an ordinal logistic regression model using the nonparametric Kruskal-Wallis test that allows for multi-factorial designs. Sex and MGS-level or sex and age were included in the regression model to reveal the cellular fractions that are significantly altered with age or MGS-level, respectively. The estimated coefficients were presented as a forest plot with 95% confidence intervals. The estimated proportions of cell types were adjusted for sex and MGS-level or sex and age for assessment of effects by factor of age or MGS-level by extracting the residuals from fitting a generalized linear model with variables.

### Statistical analyses

Detailed information of statistical analyses was described in different places, including results, figures, and figure legends, in which the test methods, significance, *p* values, error bars and coefficients were included.

### Reporting summary

Further information on research design is available in the Nature Research Reporting Summary linked to this article.

### Data availability

All processed data files generated for this study are included in this article and its supplementary information files. Raw data files from two sources are available for download through GEO under the accession number GSE115828 and Human Cell Atlas under accession number E-MTAB-7316.

### Code availability

Analysis scripts will be made available upon request.

## Data Availability

All data produced in the present study are available upon reasonable request to the authors

## Acknowledgements

We thank Ryan D. Chow for sharing codes for data analyses at Github.

## Author contribution

J-H.W. and G-S.L. conceived and designed the study. J-H.W. developed the analysis approach, performed all data analyses, and generated figures. J-H.W., R.C.B.W., and G-S.L. prepared the manuscript. G-S.L. supervised the work.

## References

1. Evans JR. Risk factors for age-related macular degeneration. Progress in retinal and eye research 20, 227–253 (2001).

2. Lambert NG, et al. Risk factors and biomarkers of age-related macular degeneration. Prog Retin Eye Res 54, 64–102 (2016).

3. Bressler NM. Age-related macular degeneration is the leading cause of blindness. JAMA 291, 1900–1901 (2004).

4. Fritsche LG, et al. A large genome-wide association study of age-related macular degeneration highlights contributions of rare and common variants. Nat Genet 48, 134–143 (2016).

5. Ratnapriya R, et al. Author Correction: Retinal transcriptome and eQTL analyses identify genes associated with age-related macular degeneration. Nat Genet 51, 1067 (2019).

6. Whitcup SM, et al. The role of the immune response in age-related macular degeneration. Int J Inflam 2013, 348092 (2013).

7. Ratnapriya R, et al. Retinal transcriptome and eQTL analyses identify genes associated with age-related macular degeneration. Nat Genet 51, 606–610 (2019).

8. Love MI, Huber W, Anders S. Moderated estimation of fold change and dispersion for RNA-seq data with DESeq2. Genome Biol 15, 550 (2014).

9. Zhou Y, et al. Metascape provides a biologist-oriented resource for the analysis of systems-level datasets. Nat Commun 10, 1523 (2019).

10. Chen M, Muckersie E, Forrester JV, Xu H. Immune activation in retinal aging: a gene expression study. Invest Ophthalmol Vis Sci 51, 5888–5896 (2010).

11. Chen M, Luo C, Zhao J, Devarajan G, Xu H. Immune regulation in the aging retina. Prog Retin Eye Res 69, 159–172 (2019).

12. Cao X, et al. Macrophage polarization in the maculae of age-related macular degeneration: a pilot study. Pathol Int 61, 528–535 (2011).

13. Kelly J, Ali Khan A, Yin J, Ferguson TA, Apte RS. Senescence regulates macrophage activation and angiogenic fate at sites of tissue injury in mice. J Clin Invest 117, 3421–3426 (2007).

14. Newman AM, et al. Determining cell type abundance and expression from bulk tissues with digital cytometry. Nat Biotechnol 37, 773–782 (2019).

15. Chen B, Khodadoust MS, Liu CL, Newman AM, Alizadeh AA. Profiling Tumor Infiltrating Immune Cells with CIBERSORT. Methods Mol Biol 1711, 243–259 (2018).

16. Tarique AA, Logan J, Thomas E, Holt PG, Sly PD, Fantino E. Phenotypic, functional, and plasticity features of classical and alternatively activated human macrophages. Am J Respir Cell Mol Biol 53, 676–688 (2015).

17. Yang Y, et al. Macrophage polarization in experimental and clinical choroidal neovascularization. Sci Rep 6, 30933 (2016).

18. Zhou Y, et al. Different distributions of M1 and M2 macrophages in a mouse model of laser-induced choroidal neovascularization. Mol Med Rep 15, 3949–3956 (2017).

19. Michetti F, et al. The S100B story: from biomarker to active factor in neural injury. J Neurochem 148, 168–187 (2019).

20. Reinehr S, et al. S100B immunization triggers NFkappaB and complement activation in an autoimmune glaucoma model. Sci Rep 8, 9821 (2018).

21. Grotegut P, et al. Minocycline reduces inflammatory response and cell death in a S100B retina degeneration model. J Neuroinflammation 17, 375 (2020).

22. Niven J, et al. S100B Up-Regulates Macrophage Production of IL1beta and CCL22 and Influences Severity of Retinal Inflammation. PLoS One 10, e0132688 (2015).

23. Ivetic A, Hoskins Green HL, Hart SJ. L-selectin: A Major Regulator of Leukocyte Adhesion, Migration and Signaling. Front Immunol 10, 1068 (2019).

24. Crane IJ, Liversidge J. Mechanisms of leukocyte migration across the blood-retina barrier. Semin Immunopathol 30, 165–177 (2008).

25. Ishimaru Y, Shibagaki F, Yamamuro A, Yoshioka Y, Maeda S. An apelin receptor antagonist prevents pathological retinal angiogenesis with ischemic retinopathy in mice. Sci Rep 7, 15062 (2017).

26. Murakami M, Simons M. Fibroblast growth factor regulation of neovascularization. Curr Opin Hematol 15, 215–220 (2008).

27. Guillonneau X, Regnier-Ricard F, Dupuis C, Courtois Y, Mascarelli F. FGF2-stimulated release of endogenous FGF1 is associated with reduced apoptosis in retinal pigmented epithelial cells. Exp Cell Res 233, 198–206 (1997).

28. Foradori MJ, et al. Matrilin-1 is an inhibitor of neovascularization. J Biol Chem 289, 14301–14309 (2014).

29. Albig AR, Schiemann WP. Fibulin-5 antagonizes vascular endothelial growth factor (VEGF) signaling and angiogenic sprouting by endothelial cells. DNA Cell Biol 23, 367–379 (2004).

30. Buga AM, Margaritescu C, Scholz CJ, Radu E, Zelenak C, Popa-Wagner A. Transcriptomics of post-stroke angiogenesis in the aged brain. Front Aging Neurosci 6, 44 (2014).

31. Shi L, et al. Newly Identified Peptide, Peptide Lv, Promotes Pathological Angiogenesis. J Am Heart Assoc 8, e013673 (2019).

32. Kurmyshkina O, Kovchur P, Schegoleva L, Volkova T. Markers of Angiogenesis, Lymphangiogenesis, and Epithelial-Mesenchymal Transition (Plasticity) in CIN and Early Invasive Carcinoma of the Cervix: Exploring Putative Molecular Mechanisms Involved in Early Tumor Invasion. Int J Mol Sci 21, (2020).

33. Cvekl A, Wang WL. Retinoic acid signaling in mammalian eye development. Exp Eye Res 89, 280–291 (2009).

34. Pollock LM, Xie J, Bell BA, Anand-Apte B. Retinoic acid signaling is essential for maintenance of the blood-retinal barrier. FASEB J 32, 5674–5684 (2018).

35. da Silva S, Cepko CL. Fgf8 Expression and Degradation of Retinoic Acid Are Required for Patterning a High-Acuity Area in the Retina. Dev Cell 42, 68–81 e66 (2017).

36. Takeuchi H, Yokota A, Ohoka Y, Iwata M. Cyp26b1 regulates retinoic acid-dependent signals in T cells and its expression is inhibited by transforming growth factor-beta. PLoS One 6, e16089 (2011).

37. Zolfaghari R, Chen Q, Ross AC. DHRS3, a retinal reductase, is differentially regulated by retinoic acid and lipopolysaccharide-induced inflammation in THP-1 cells and rat liver. Am J Physiol Gastrointest Liver Physiol 303, G578–588 (2012).

38. Fleckenstein M, et al. The Progression of Geographic Atrophy Secondary to Age-Related Macular Degeneration. Ophthalmology 125, 369–390 (2018).

39. Deutschman CS, Tracey KJ. Sepsis: current dogma and new perspectives. Immunity 40, 463–475 (2014).

40. Tracey KJ. Reflexes in Immunity. Cell 164, 343–344 (2016).

41. Pavlov VA, Tracey KJ. Neural regulation of immunity: molecular mechanisms and clinical translation. Nat Neurosci 20, 156–166 (2017).

42. Stofkova A, Kamimura D, Ohki T, Ota M, Arima Y, Murakami M. Photopic light-mediated down-regulation of local alpha1A-adrenergic signaling protects blood-retina barrier in experimental autoimmune uveoretinitis. Sci Rep 9, 2353 (2019).

43. London A, Benhar I, Schwartz M. The retina as a window to the brain-from eye research to CNS disorders. Nat Rev Neurol 9, 44–53 (2013).

44. Omary MB, Coulombe PA, McLean WH. Intermediate filament proteins and their associated diseases. N Engl J Med 351, 2087–2100 (2004).

45. Wu KH, Madigan MC, Billson FA, Penfold PL. Differential expression of GFAP in early v late AMD: a quantitative analysis. Br J Ophthalmol 87, 1159–1166 (2003).

46. Daugan MV, et al. Complement C1s and C4d as Prognostic Biomarkers in Renal Cancer: Emergence of Noncanonical Functions of C1s. Cancer Immunol Res 9, 891–908 (2021).

47. Newman AM, et al. Systems-level analysis of age-related macular degeneration reveals global biomarkers and phenotype-specific functional networks. Genome Med 4, 16 (2012).

48. Natoli R, et al. Retinal Macrophages Synthesize C3 and Activate Complement in AMD and in Models of Focal Retinal Degeneration. Invest Ophthalmol Vis Sci 58, 2977–2990 (2017).

49. Gold MC, Napier RJ, Lewinsohn DM. MR1-restricted mucosal associated invariant T (MAIT) cells in the immune response to Mycobacterium tuberculosis. Immunol Rev 264, 154–166 (2015).

50. Fan W, Li X, Wang W, Mo JS, Kaplan H, Cooper NG. Early Involvement of Immune/Inflammatory Response Genes in Retinal Degeneration in DBA/2J Mice. Ophthalmol Eye Dis 1, 23–41 (2010).

51. Patak J, Hess JL, Zhang-James Y, Glatt SJ, Faraone SV. SLC9A9 Co-expression modules in autism-associated brain regions. Autism Res 10, 414–429 (2017).

52. Lukowski SW, et al. A single-cell transcriptome atlas of the adult human retina. EMBO J 38, e100811 (2019).

53. Menon M, et al. Single-cell transcriptomic atlas of the human retina identifies cell types associated with age-related macular degeneration. Nat Commun 10, 4902 (2019).

54. Vecino E, Rodriguez FD, Ruzafa N, Pereiro X, Sharma SC. Glia-neuron interactions in the mammalian retina. Prog Retin Eye Res 51, 1–40 (2016).

55. Reichenbach A, Bringmann A. New functions of Muller cells. Glia 61, 651–678 (2013).

56. Ma W, Wong WT. Aging Changes in Retinal Microglia and their Relevance to Age-related Retinal Disease. Adv Exp Med Biol 854, 73–78 (2016).

57. Anderson DH, et al. The pivotal role of the complement system in aging and age-related macular degeneration: hypothesis re-visited. Prog Retin Eye Res 29, 95–112 (2010).

58. Dinu V, Miller PL, Zhao H. Evidence for association between multiple complement pathway genes and AMD. Genet Epidemiol 31, 224–237 (2007).

59. Ding X, Patel M, Chan CC. Molecular pathology of age-related macular degeneration. Prog Retin Eye Res 28, 1–18 (2009).

60. Waksmunski AR, et al. Pathway Analysis Integrating Genome-Wide and Functional Data Identifies PLCG2 as a Candidate Gene for Age-Related Macular Degeneration. Invest Ophthalmol Vis Sci 60, 4041–4051 (2019).

61. Jones BW, Pfeiffer RL, Ferrell WD, Watt CB, Tucker J, Marc RE. Retinal Remodeling and Metabolic Alterations in Human AMD. Front Cell Neurosci 10, 103 (2016).

62. Curcio CA, Medeiros NE, Millican CL. Photoreceptor loss in age-related macular degeneration. Invest Ophthalmol Vis Sci 37, 1236–1249 (1996).

63. Curcio CA. Photoreceptor topography in ageing and age-related maculopathy. Eye (Lond) 15, 376–383 (2001)

64. Sonntag S, et al. Ablation of retinal horizontal cells from adult mice leads to rod degeneration and remodeling in the outer retina. J Neurosci 32, 10713–10724 (2012).

65. Yi W, et al. A single-cell transcriptome atlas of the aging human and macaque retina. Natl Sci Rev 8, nwaa179 (2021).

66. Johnson PT, et al. Drusen-associated degeneration in the retina. Invest Ophthalmol Vis Sci 44, 4481–4488 (2003).

67. Lyu Y, et al. Implication of specific retinal cell-type involvement and gene expression changes in AMD progression using integrative analysis of single-cell and bulk RNA-seq profiling. Sci Rep 11, 15612 (2021).

68. McLeod DS, Grebe R, Bhutto I, Merges C, Baba T, Lutty GA. Relationship between RPE and choriocapillaris in age-related macular degeneration. Invest Ophthalmol Vis Sci 50, 4982–4991 (2009).

69. Stuart T, et al. Comprehensive Integration of Single-Cell Data. Cell 177, 1888–1902 e1821 (2019).

70. Wadi L, Meyer M, Weiser J, Stein LD, Reimand J. Impact of outdated gene annotations on pathway enrichment analysis. Nat Methods 13, 705–706 (2016).

71. Liu Q, Shepherd BE, Li C, Harrell FE, Jr. Modeling continuous response variables using ordinal regression. Stat Med 36, 4316–4335 (2017).

72. Ritchie ME, et al. limma powers differential expression analyses for RNA-sequencing and microarray studies. Nucleic Acids Res 43, e47 (2015).

73. Wei T, Simko V, Levy M, Xie Y, Jin Y, Zemla J. corrplot: Visualization of a correlation matrix. R package version 073 230, (2013).

74. Chow RD, Majety M, Chen S. The aging transcriptome and cellular landscape of the human lung in relation to SARS-CoV-2. Nat Commun 12, 4 (2021).

75. Newman AM, et al. Robust enumeration of cell subsets from tissue expression profiles. Nat Methods 12, 453–457 (2015).

